# Starting, stopping and restarting. Patterns of Methylphenidate Use over 14 years in a large public health system

**DOI:** 10.64898/2026.06.18.26355862

**Authors:** Mark Richards, Hendrike McDonald, Veruschka Ramanjam, Estelle Lawrence, Kirsten Donald

**Affiliations:** Department of Paediatrics and Child Health, Faculty of Health Sciences, University of Cape Town, Cape Town, South Africa; Western Cape Department of Health and Wellness, Cape Town, South Africa

## Abstract

**Background:** Persistence with stimulant medication is poor in children and adolescents with ADHD, and the evidence base is derived predominantly from high-income countries. We describe methylphenidate utilisation patterns and predictors of 12-month retention across 14 years in a large South African public health service.

**Methods:** Retrospective cohort study using routine pharmacy data from the Western Cape provincial health service (2011-2024). Children aged 5-18 at first prescription were included. Treatment episodes were defined as continuous prescription sequences with no gap exceeding 90 days and classified as initiations or restarts. Logistic regression modelled 12-month retention against early visit frequency and formulation type as pre-specified exposures.

**Findings:** 421,925 prescription events for 23,243 children across 115 facilities generated 65,885 treatment episodes. Median age at first prescription was 10 years (IQR 8-12); 77·6% were male. Kaplan-Meier 12-month survival was 28·2% for initiations and 15·4% for restarts, substantially below high-income country comparators. A quarter of all initiating prescriptions were not followed by a subsequent dispensing event; nearly 40% of patients had three or more treatment episodes. Early visit frequency was the strongest predictor of 12-month retention (high vs low: OR 2·85, 95% CI 2·65-3·06). The sustained-release formulation effect was present but attenuated on multivariable adjustment. Treatment re-initiations showed a marked seasonal pattern consistent with the South African school calendar.

**Interpretation:** Twelve-month retention was markedly lower than high-income country rates. Against a backdrop of high attrition, both early visit frequency and sustained-release formulation access predicted persistence; clinical engagement and reducing structural barriers to access are modifiable factors in this setting.

**Funding:** None.

## Introduction

Attention-deficit/hyperactivity disorder (ADHD) is a common and chronic neurodevelopmental condition, affecting approximately 3.4% of children and adolescents globally^1, 2^. It is associated with deleterious effects on learning, scholastic outcomes and personal and social wellbeing, compounded in low- and middle-income countries (LMIC) by limited access to diagnosis and capacitated therapeutic ecosystems^1, 3^. The positive effects of stimulant medication on mitigating these outcomes, are well attested in published literature. Long term persistence has reported rates of between 50% and 65% at one year, dropping to approximately 20% at five years^4–7^. A recent systematic review article by Ferrin et al. (2025), established from their pooled data that only 22·9% of children and adolescents had good adherence at 12 month follow up^8^. This is consistent with systematic reviews from earlier literature^7, 9^.

The essential quality of engagement between health services and patients and families in long term neurobehavioural conditions is foundational and has a strong published support^10–12^. Within ADHD specifically, a number of service descriptions and national guidelines are at lengths to define the elements of a caring and attentive service, mindful of the challenges inherent in the long-term management of this condition^13–18^. However, published evidence on verifying interventions to enhance long term regular medication use is not well developed^8, 19,20^. Predictors of persistence are varied and, depending on the methodology, focus on several service, parent and patient variables ^21–25^.

Research in the field of adherence and persistence struggles with a lack of consensus on definitions and methodologies, in particular, duration thresholds and the maximum duration between treatment episodes^8^. Compliance and adherence, often used interchangeably, imply following instructions from a healthcare provider. Persistence specifically measures the duration of continuous treatment from initiation to cessation, addressing a temporal dimension^26^. This article will focus primarily on persistence.

ADHD and its treatment in general, and use patterns of stimulants in particular, have a comparatively small literature from LMICs as a category^3, 27^. The condition is described and access to medication is limited, but relatively little is known about how this medication is used in resource limited settings. In the review article by Ferrin et al., (cited above), only 0·3% of the patients described in 66 articles were from LMIC’s^8^. South Africa has a small literature on the subject, though predominantly from private healthcare settings. Key findings have been the challenges of access for children with ADHD, misuse amongst students without ADHD for performance enhancement, and seasonal use patterns coinciding with high assessment intensity^28–32^. There is a limited literature on longitudinal use patterns, and this with small patient numbers, from other LMIC’s^33–35^.

In South Africa, the restricted scheduling status of stimulants prohibits prescription of volumes of medication that exceed daily use beyond 30 days, with all subsequent prescriptions requiring physical re-writes by a medical practitioner. Attendance at a health facility is required, often taking several hours of waiting for a doctor to write a prescription and receive the medication at a facility pharmacy.

Public health services in the Western Cape Province (population 7·43 million) operate under severe resource constraints, with limited access to professionals with child mental health expertise^36^. The communities that are served are also characterised by high levels of poverty and violence and many schools have severely overcrowded classrooms -conditions that can mimic and exacerbate ADHD symptomatology^37–39^. Recognition of mental health conditions as treatable is inconsistent, and engagement with mental health services is constrained by structural barriers such as poverty and vulnerable employment status, and alternative explanatory models of a childhood behavioural condition ^40, 41^. These factors frame the treatment ecosystem within which our cohort was defined.

The data in this article describe an electronic data repository of prescription data for methylphenidate for the multiple dispensing sites of the provincial state health service in the Western Cape Province of South Africa (population 7·4 million). In this study we describe the patterns and durations of methylphenidate prescriptions for children under 18 at the commencement of their care in the Western Cape public health service and explore predictors of long-term treatment persistence.

## Methods

This was a retrospective cohort study using routinely collected pharmacy data in the state health service in the Western Cape Province of South Africa from the beginning of 2011 to the end of 2024. The majority of the medically uninsured province’s population will receive care in this service. Children and adolescents from 5 to 18 years of age at first recorded prescription date were included in the study. Prescription data were accessed from the Western Cape Department of Health Information Directorate for every methylphenidate prescription issued to individuals over the study period. Institutions within the Western Cape progressively contributed to the dataset from 2011, initially for large hospitals and then progressively including smaller day clinics over subsequent years.

A prescription event corresponded to a clinical encounter on a single calendar day. Row level data did not include diagnostic information nor daily dosage details and pill volume records were too irregular to allow dosing extrapolation. Formulations were designated as either Immediate Release (IR), Sustained Release (SR), or Both -- where the two formulations were dispensed on the same day. Each prescription event was assigned a duration of one month, reflecting the legal maximum 30-day prescription period for stimulant medication in South Africa. Formulation type was recorded discretely for each event and used in formulation analyses. (See Figure 1).

**Figure 1.**
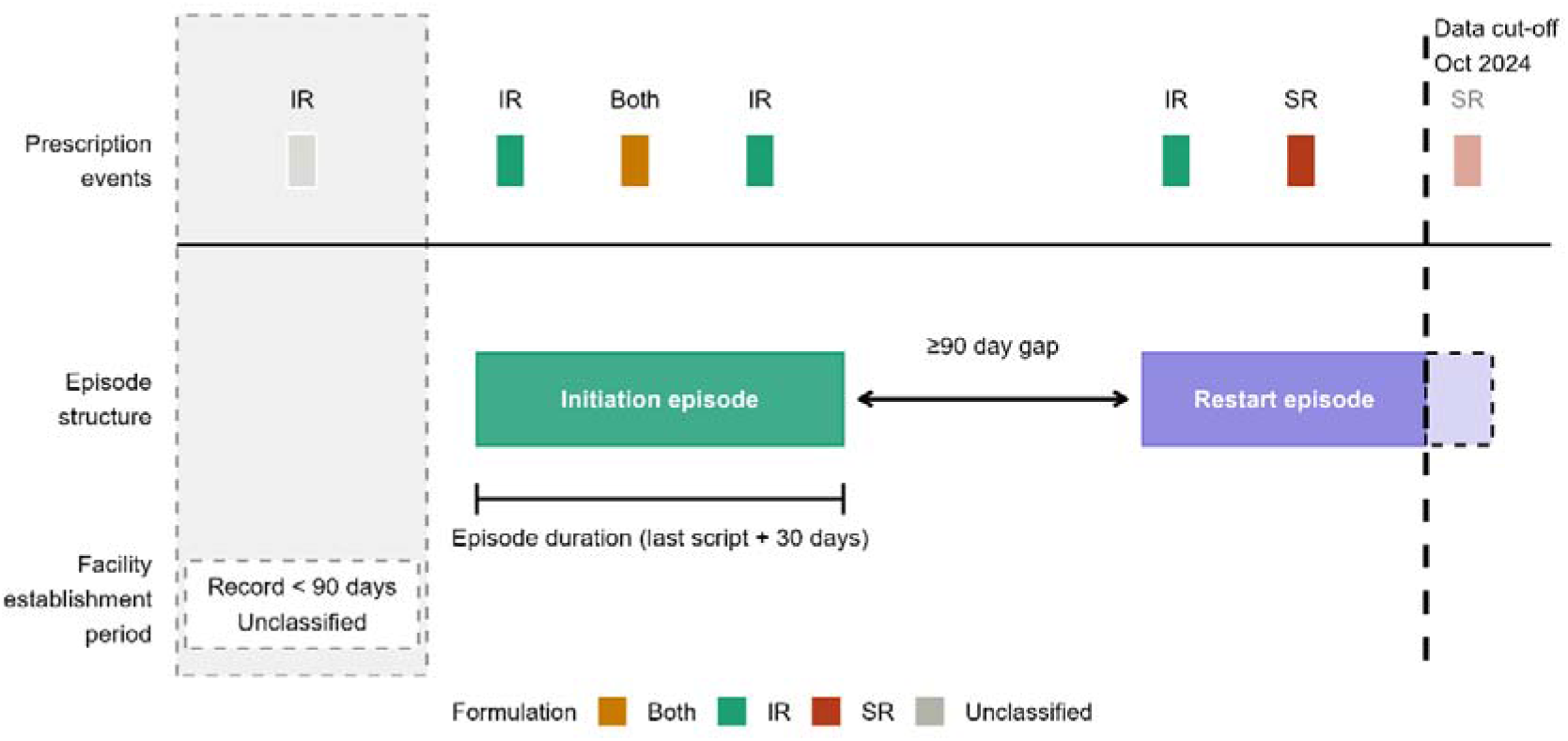
Prescriptions and treatment episodes visual representation

A treatment episode is a continuous period of treatment defined as an unbroken sequence of prescription events where no gap between consecutive events exceeded 90 days. An episode was considered closed when 90 or more days elapsed without a further prescription event. Episode duration was calculated from the first prescription event to 30 days after the final prescription event, with the additional 30 days reflecting the medication supply issued on the last date.

An episode was classified as an initiation if it represented the patient’s first recorded treatment episode at an established facility. A restart was defined as any episode where the preceding treatment episode had ended more than 90 days before the first prescription event of the new episode. To allow reliable episode differentiation, a facility establishment period of 90 days from the date of the first recorded prescription at any given facility was applied. Prescription events falling within this window could not be classified with certainty as new initiations or continuations of prior treatment and were therefore excluded from the primary analyses. These were designated as unclassified.

Transfers between facilities were recorded but did not affect individual episode duration calculations. Treatment episodes containing prescription events within the final three months of 2024 were right-censored for survival analysis, meaning the episode was still ongoing at the point data collection ended and its true duration therefore remains unknown.

Patients were categorised by gender as recorded in the pharmacy data; records where gender was unassigned or inconsistent were noted but excluded from gender-specific analyses. Age at first prescription was used to assign patients to three groups: 5-9 years, 10-14 years, and 15-18 years.

The study period was marked by the system-level loss of access to SR methylphenidate in 2022, consequent to escalating costs related to price increases from suppliers, an event previously described in our setting in 2008^42^. After this, until the closure of the study period, only IR methylphenidate was available in the state service.

Analysis of the associations with visit frequency and formulation type was restricted to episodes longer than three months. Doctors generally assign review intervals and so visit frequency in the first three months was used as a proxy for the intensity of clinical engagement between doctors and families. Episodes with a shorter review period than the standard four weeks were categorised as high visit frequency (>1·1 visits/month), those consistent with standard monthly prescribing as medium frequency (0·9-1·1 visits/month), and those with intervals longer than the regulated 30-day prescription period as low frequency (<0·9 visits/month).

Descriptive statistics employed non-parametric methods throughout. Kaplan-Meier survival analysis was stratified by episode type and early treatment visit frequency (first three months). Episodes ongoing at the data cut-off were included as censored observations.

Episodes at facilities with insufficient prescribing history (less than 90 days of records at the time of the patient’s first script) were excluded from survival analyses but retained in descriptive analyses. As patients could contribute multiple restart episodes, these were treated as independent observations.

Logistic regression was used to model 12-month treatment retention. Univariable models were first fitted for visit frequency category and formulation type separately. Both variables were then entered simultaneously into a multivariable model, selected *a priori* as the two primary exposures of interest, to examine the independent contribution of each after mutual adjustment.

The statistical package R (Posit Software, PBC, version 4·5.0, 2025-04-11 ucrt using RStudio 2025·09·1) was used for data cleaning, analysis and graphic generation, with assistance from Claude Sonnet 4·6 (Anthropic, PBC, accessed 9 February 2026) for code generation. The authors take full responsibility for the analysis.

Ethical approval for this study was granted by the University of Cape Town Faculty of Health Sciences Human Research Ethics Committee (HREC Ref: 861/2023). A waiver of informed consent was granted in view of the retrospective nature of the study and the size of the cohort.

## Results

Over the 14 years of the study period there were 421,925 prescription events of methylphenidate formulations for 23,243 children at 115 facilities documented on the Western Cape Health Department’s electronic prescription database. The vast majority (72·4%) of prescription events were for IR methylphenidate formulations only. Most of these prescriptions were for males (77·6%), and initiations were most frequent for children under 10 years of age, with a peak at 7-8 years of age coinciding with the 2^nd^ and 3rd year of formal schooling. Subsequent treatment episodes demonstrated a shift to the older child (Fig.2) with a peak towards the end of primary school and the start of secondary school when academic demands tend to escalate.

**Figure 2.**
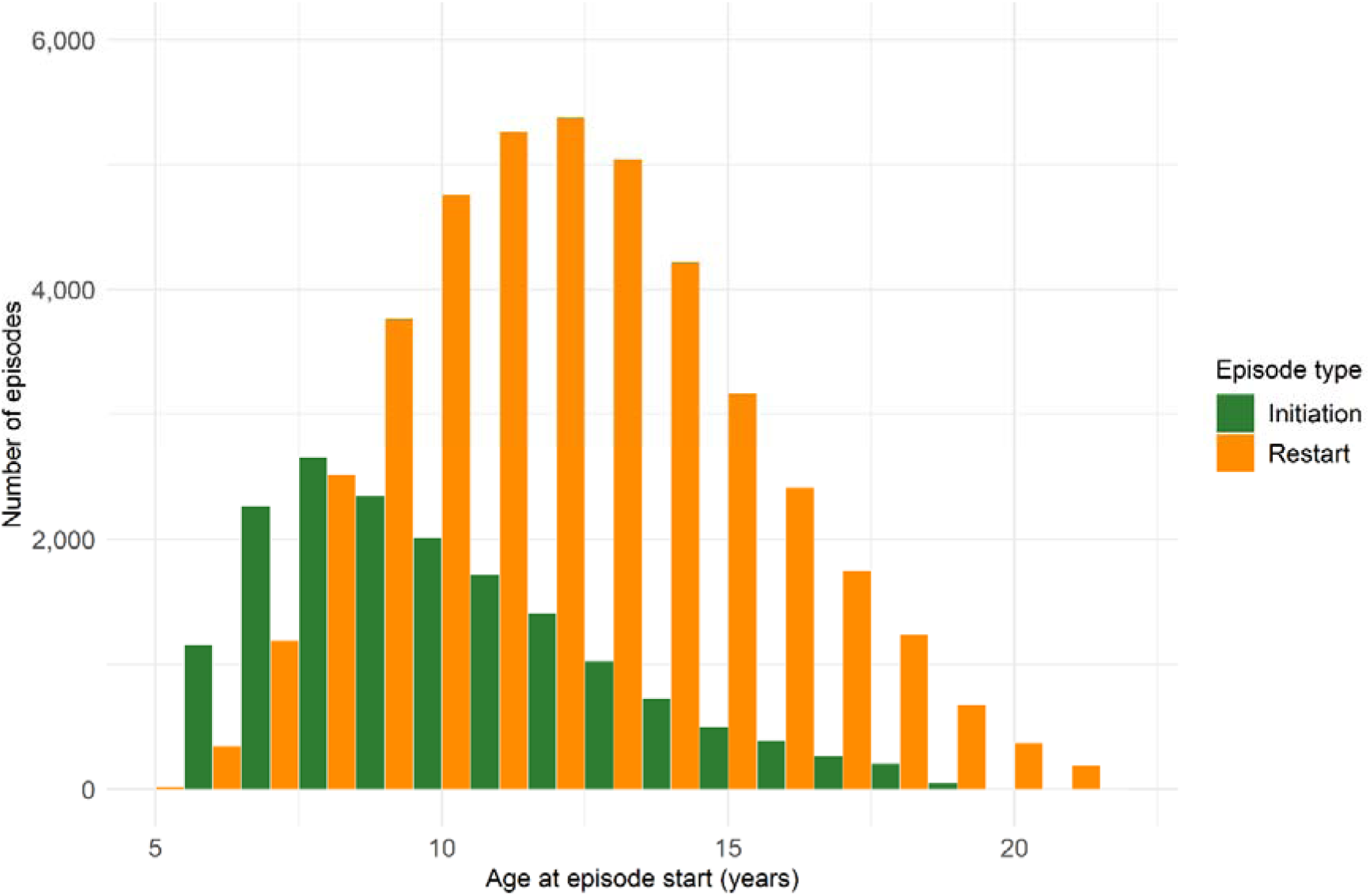
Age distribution at episode start, by episode type. Western Cape public health services, 2011-2024. Note: 4·4% of patients contributed episodes beyond 18 years of age, having initiated treatment before they were 18 and then remained till study period closure.

Among all patients, 41·2% of patients had a single treatment episode and 39·6% of patients had three or more distinct treatment episodes. Of initation episodes, 25·4% comprised only a single script, and of re-initiations, 29·7% had only a single script. Individuals tended to remain at their initiating facility with only a small number of patients moving between facilities (8·3%). It was not possible to discern transitions between clinicians within an institution for an individual child or adolescent.

Crude retention proportions to 12 months were 20·5% for first initiations and 11·1% for restarts respectively. Attrition rates were highest in the first few months with approximately half of all episodes ending 3 -4 months after starting; single-script episodes alone accounted for nearly a quarter of episodes. Kaplan-Meier 12-month survival estimates are 28·2% and 15·4% respectively, with the difference between episode types consistent across time (Fig. 3).

**Figure 3.**
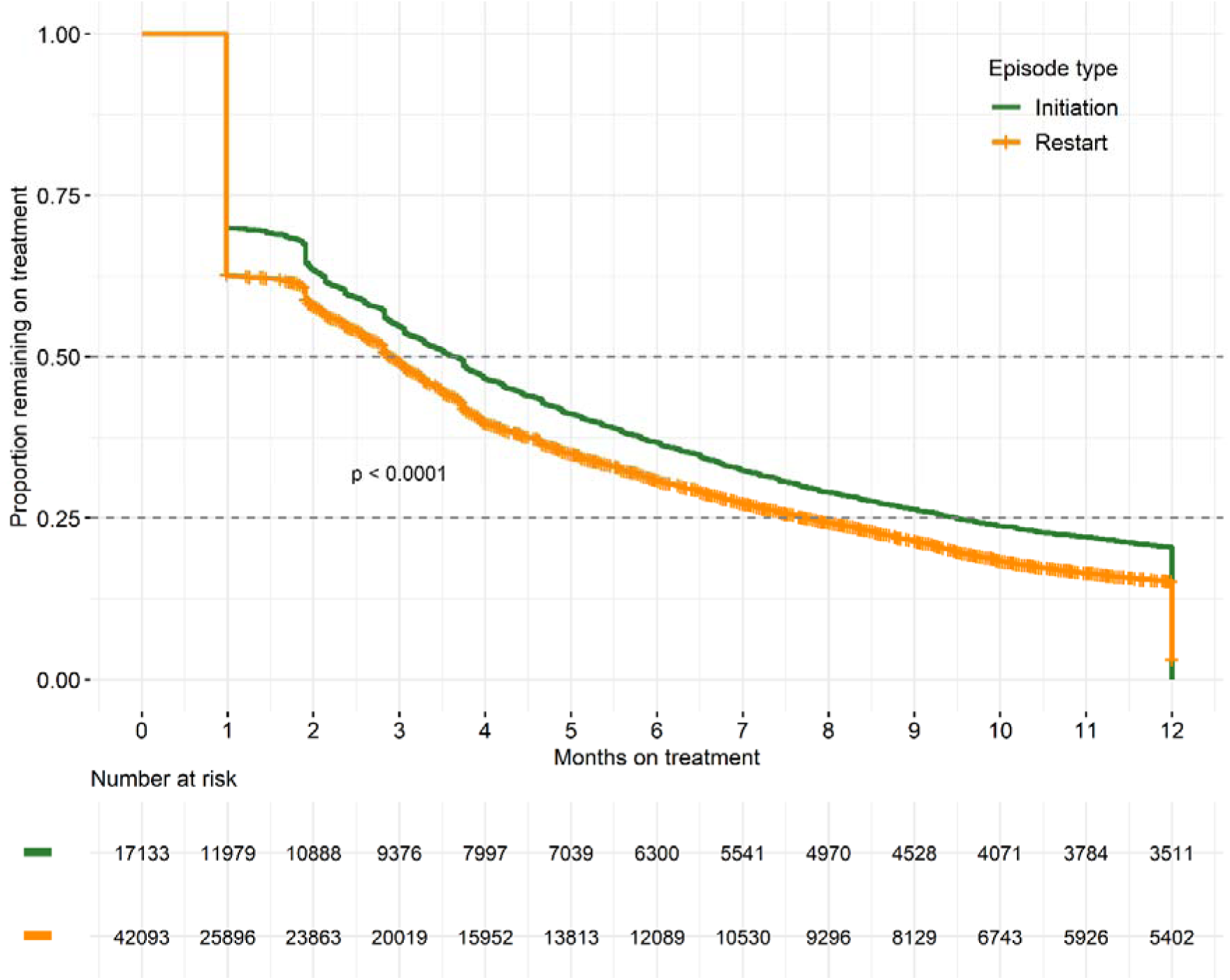
Kaplan-Meier survival curves by episode type. Western Cape public health services, 2011-2024.

Survival curves demonstrate consistent separation by early visit frequency category (Fig. 4) with a steadily increasing likelihood for long term retention of patient care. 53·4% of high-frequency episodes remained on treatment at 12 months compared with 38·0% for medium and 25·7% for low frequency, with survival widening over time (Fig 4, p<0·0001).

**Figure 4.**
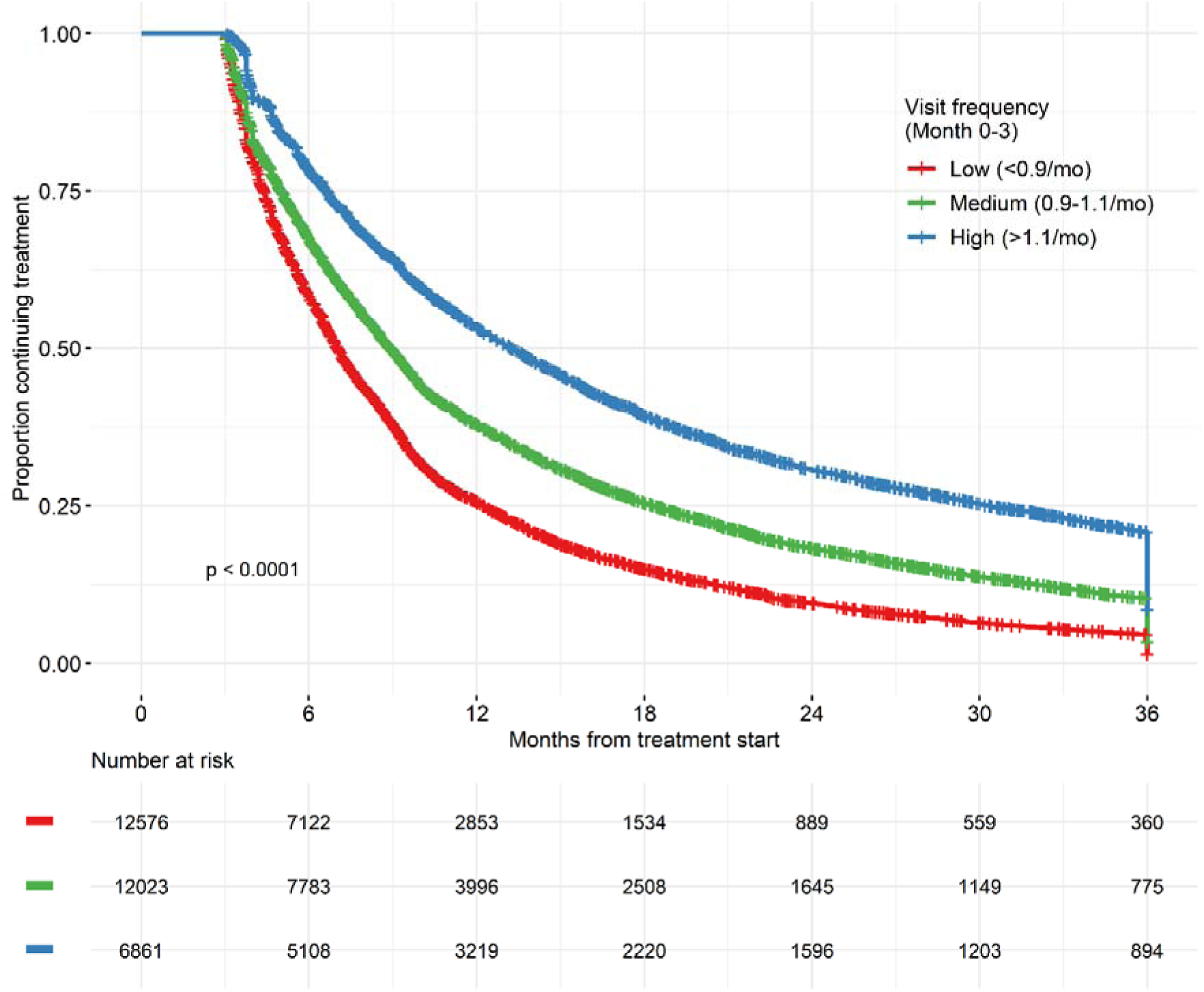
Kaplan-Meier survival curves by early visit frequency. Western Cape public health services, 2011-2024.

Episode duration did not significantly differ by gender, but age group episodes had progressively shorter treatment durations as children got older. Facility transitions (7·5% of episodes) appeared to confer an advantage, we surmise by improved access to a more proximate facility.

We hypothesised that early visit frequency and a mixed formulation use would be indicators of higher engagement and attentive medical care, essentially defining a “positive deviants” cohort^43^. The former variable indicating a shorter follow-up interval than a standard prescription period of 30 days and the latter an indicator of possible dose titration from IR to an established SR dose. For this we employed an a priori logistic regression rather than a stepwise method, to understand their predictive ability for 12 month retention. When the first three months of this cohort of patients was isolated as a period of interest, both high visit frequency (seeing a doctor more than once a month) and the use of an SR formulation, independently predicted 12 month retention using a univariable model. There was consistently stronger predictive value for each of the categories of higher visit frequency.

In a multivariable model, higher visit frequency, for both medium and high categories, remained a significant predictor of 12-month retention (Table 4) and visit frequency ORs were essentially unchanged. Although still significant, the effect of stimulant formulations was attenuated - the “Both (IR+SR)” Odds Ratio (OR) dropped from 1·42 to 1·28, and “SR only” increased slightly from 1·20 to 1·24.

**Table 1.**
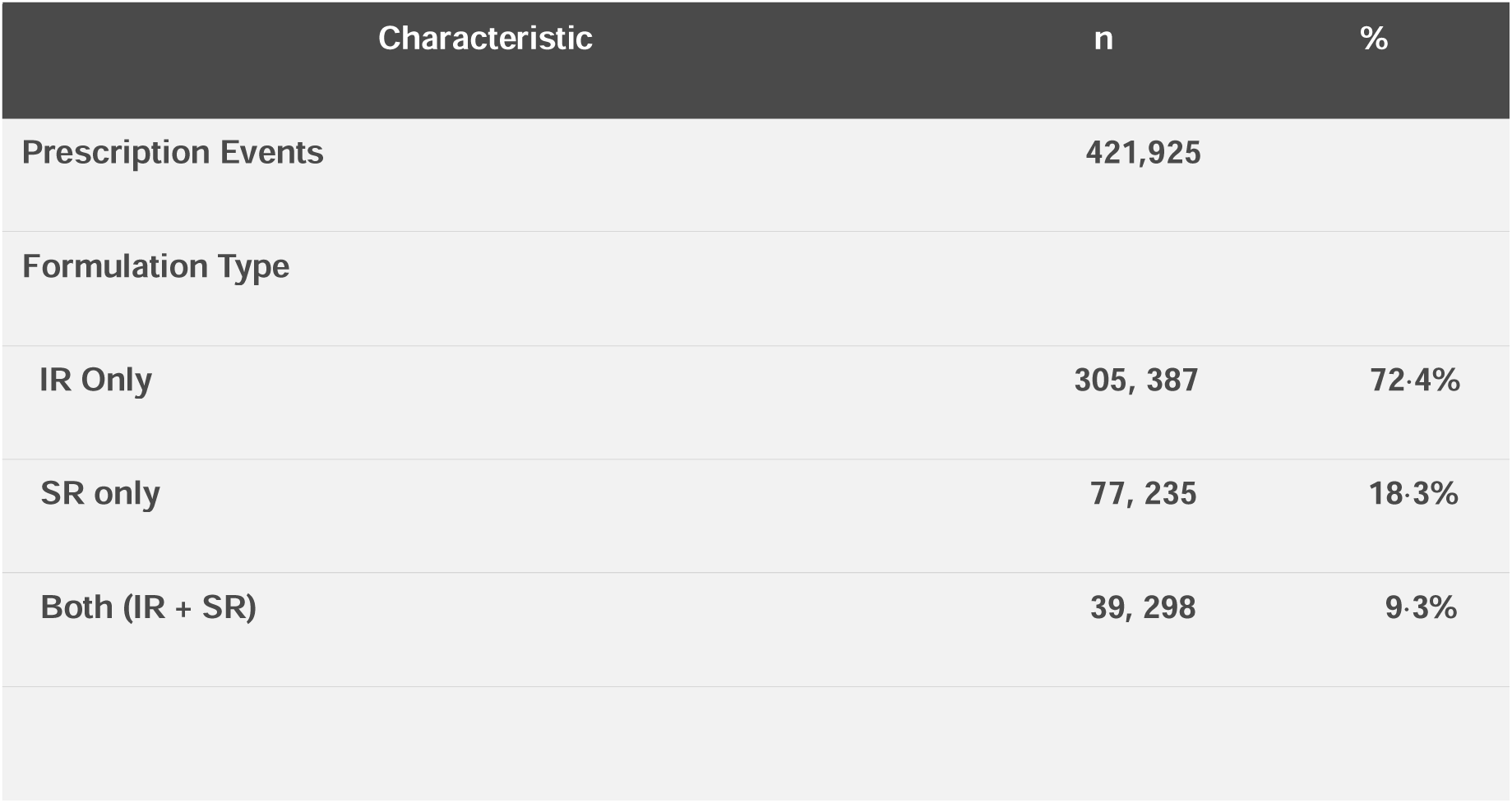
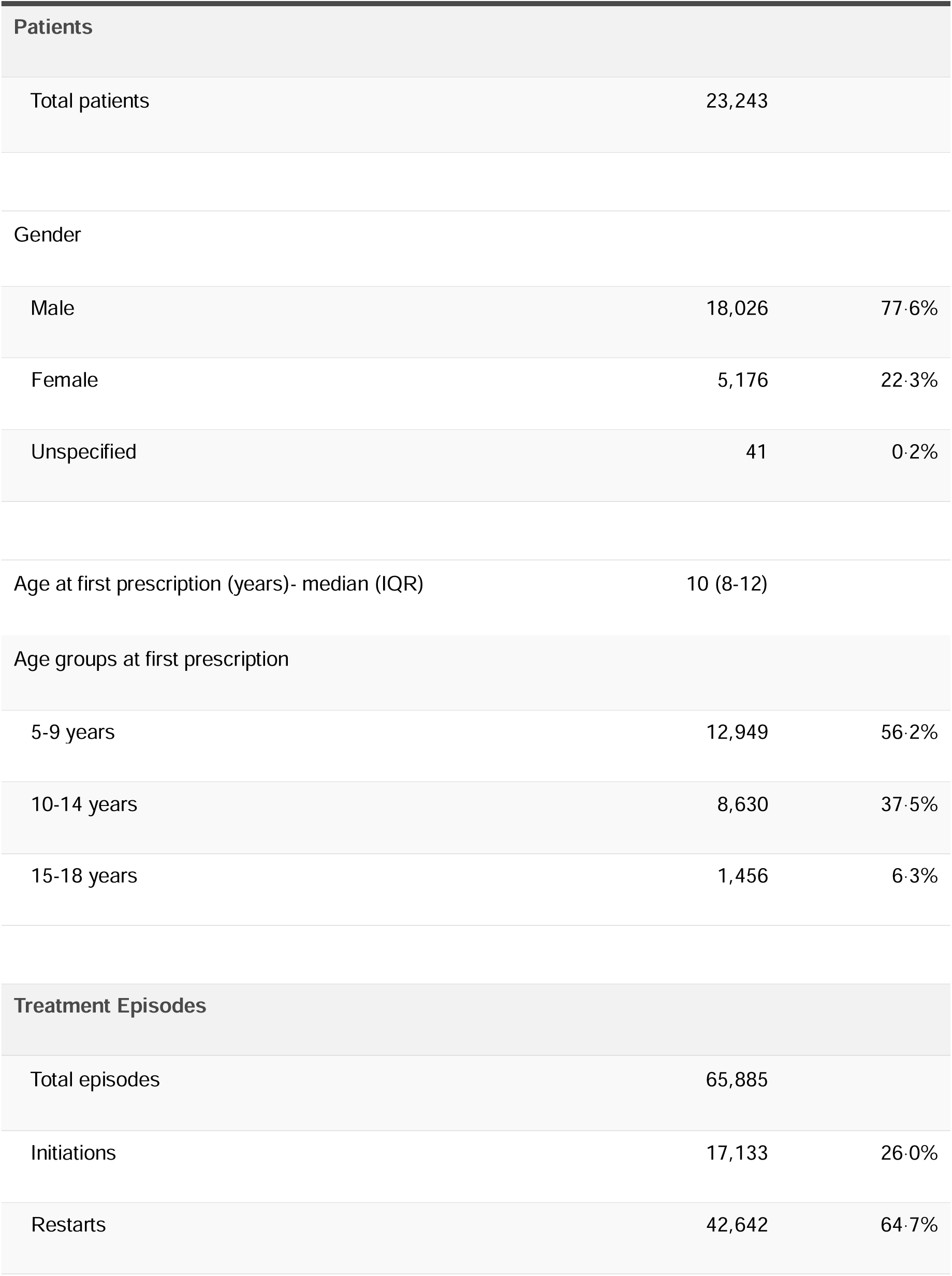
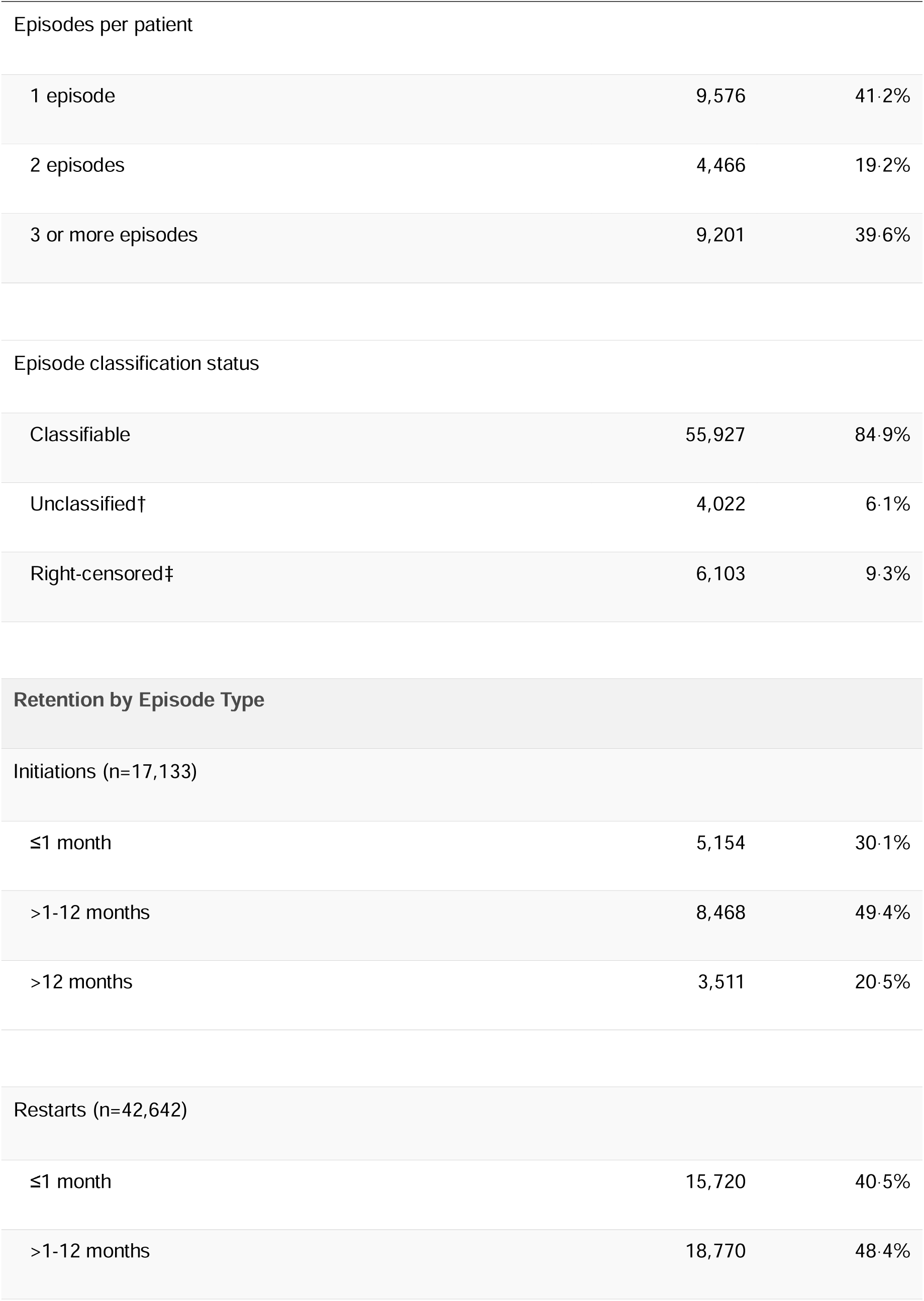
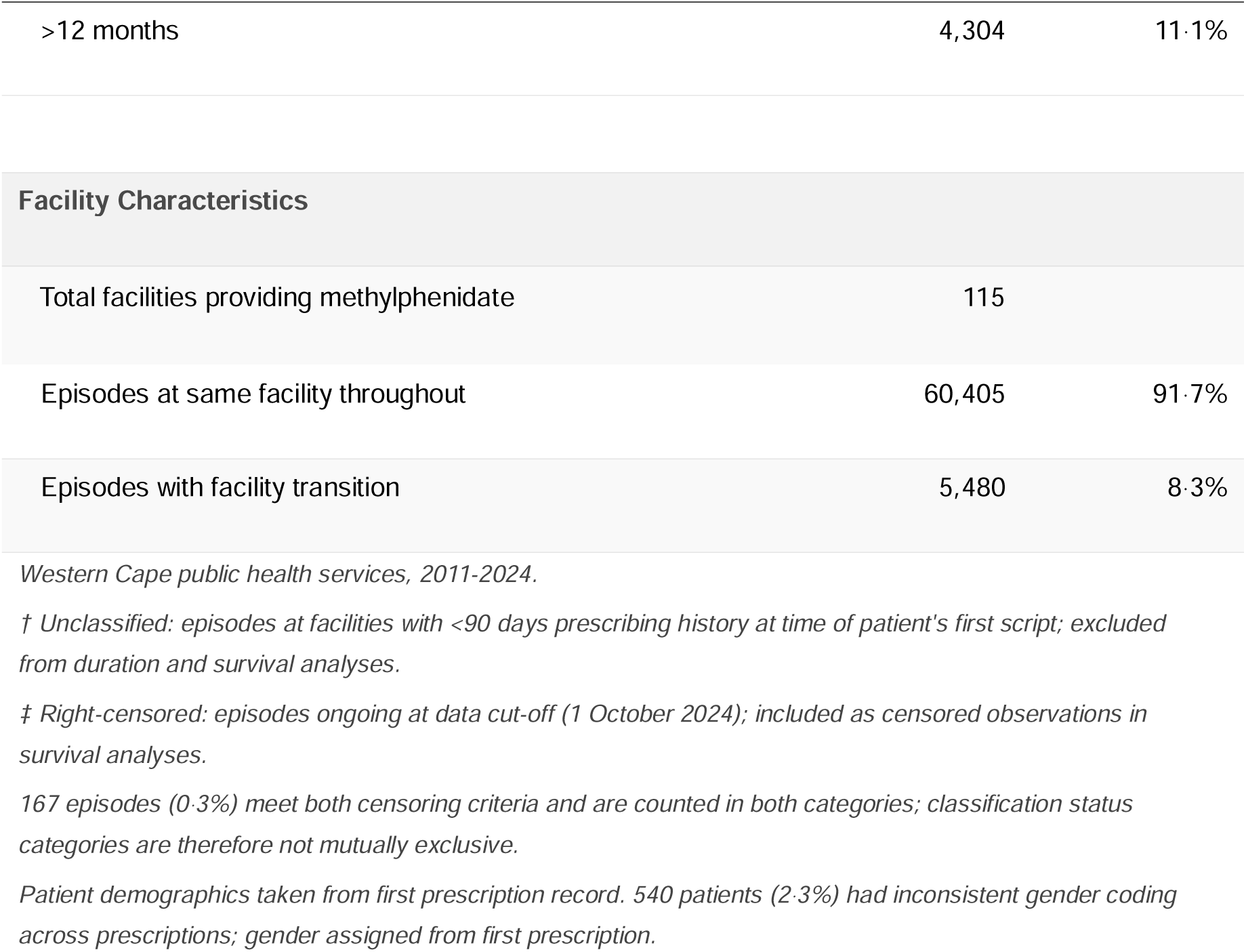
Study population and treatment episode characteristics.

**Table 2.** Treatment episode durations by patient and episode characteristics.

| Variable | Category | n | Median (IQR) |
| --- | --- | --- | --- |
| Episode Type | Initiation | 17,133 | 3.6 (1.0-9.5) |
|  | Restart | 38,794 | 2.6 (1.0-6.4) |
| Gender | Female | 12,185 | 2.9 (1.0-7.3) |
|  | Male | 43,651 | 2.8 (1.0-7.2) |
|  | Unspecified | 91 | 3.3 (1.0-7.0) |
| Age Group | 5-9 years | 18,981 | 3.1 (1.0-8.6) |
|  | 10-14 years | 28,184 | 2.8 (1.0-6.9) |
|  | 15-18 years | 6,787 | 2.5 (1.0-6.1) |
|  | 19+ years** | 1,975 | 2.1 (1.0-5.6) |
| Facility Transition<sup>†</sup> | None | 50,174 | 2.3 (1.0-6.1) |
|  | Permanent Transfer | 4,210 | 8.4 (3.8-18.7) |
|  | Temporary Transfer | 1,543 | 10.4 (6.5-20.8) |
| Formulation (Month 0-3)* | IR only | 18,333 | 7.2 (4.5-12.8) |
|  | SR only | 4,411 | 8.0 (4.7-14.7) |
|  | Both (IR+SR) | 4,201 | 8.2 (4.8-16.2) |
| Visit Frequency (Month 0-3) * | Low (<0.9/mo) | 11,372 | 6.5 (4.2-10.4) |
|  | Medium (0.9-1.1/mo) | 10,256 | 7.9 (4.7-14.3) |
|  | High (>1.1/mo) | 5,317 | 10.2 (5.7-19.4) |

| Variable | Category | n | Median (IQR) |
| --- | --- | --- | --- |

**Table 3.** Twelve-month treatment retention by formulation type and early visit frequency.

| Formulation (Month 0-3) | Low (<0.9/mo) | Medium (0.9-1.1/mo) | High (>1.1/mo) |
| --- | --- | --- | --- |
| IR only | 20.2% (1,615/8,007) | 29.1% (2,016/6,932) | 40.0% (1,359/3,394) |
| SR only | 22.2% (448/2,017) | 33.8% (557/1,649) | 48.9% (364/745) |
| Both (IR+SR) | 22.5% (303/1,348) | 34.6% (580/1,675) | 48.6% (573/1,178) |

Values are % retained >12 months (n retained / total n).
Episodes $\geq 3$ months, classifiable only (N=26,945).
Visit frequency defined as scripting events per month during the first 3 months of the episode.

**Table 4.**
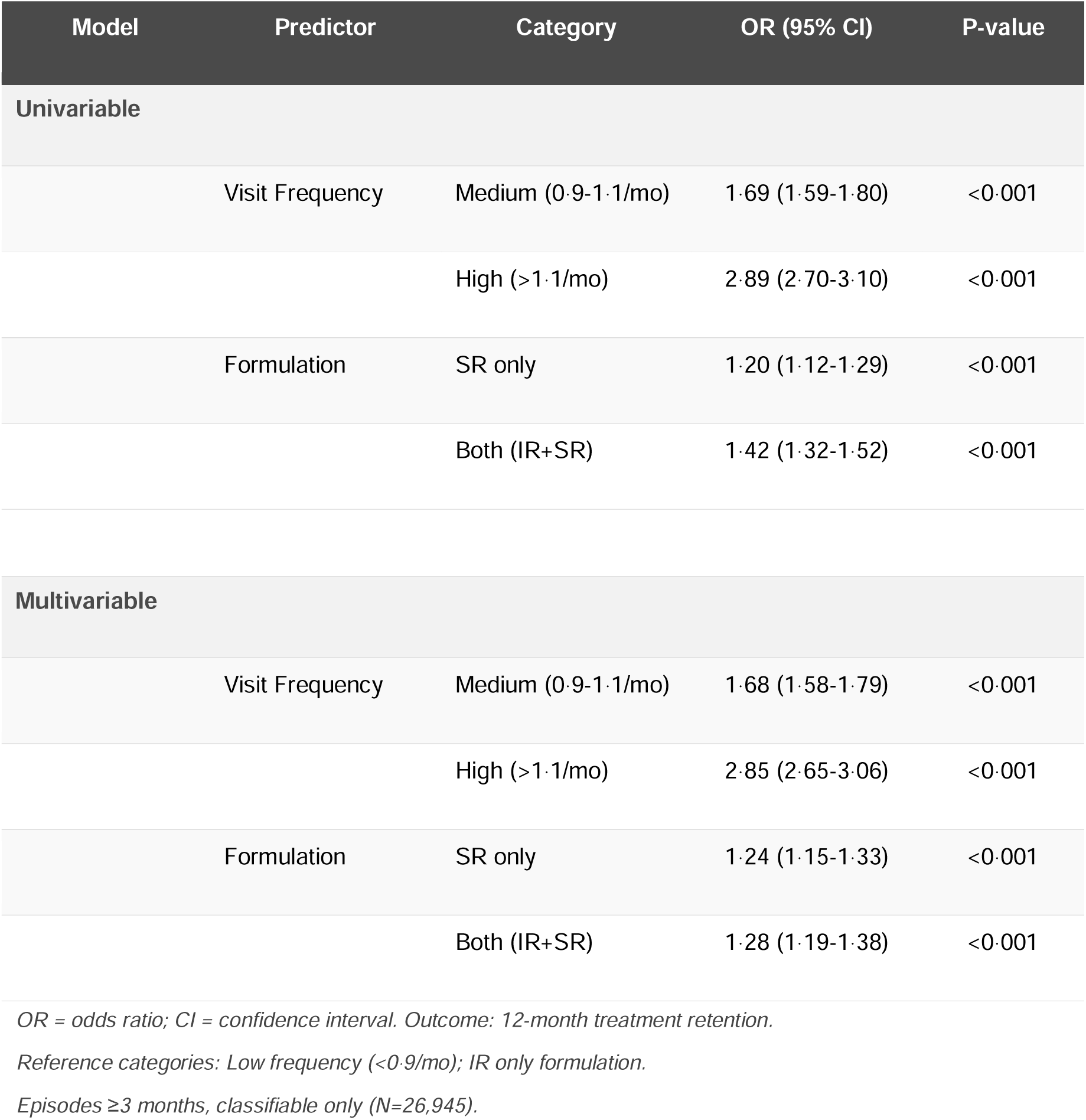
Logistic regression: predictors of 12-month treatment retention.

Prescription events by calendar month, categorised by either initiation or restart episodes, show some similarity to each other in temporal patterns (Figure 5). Initiations show some peaking in the months prior to mid and end of year exams. There is a marked peak in the re-initiation of treatment after the mid-year and end of year vacations.

**Figure 5.**
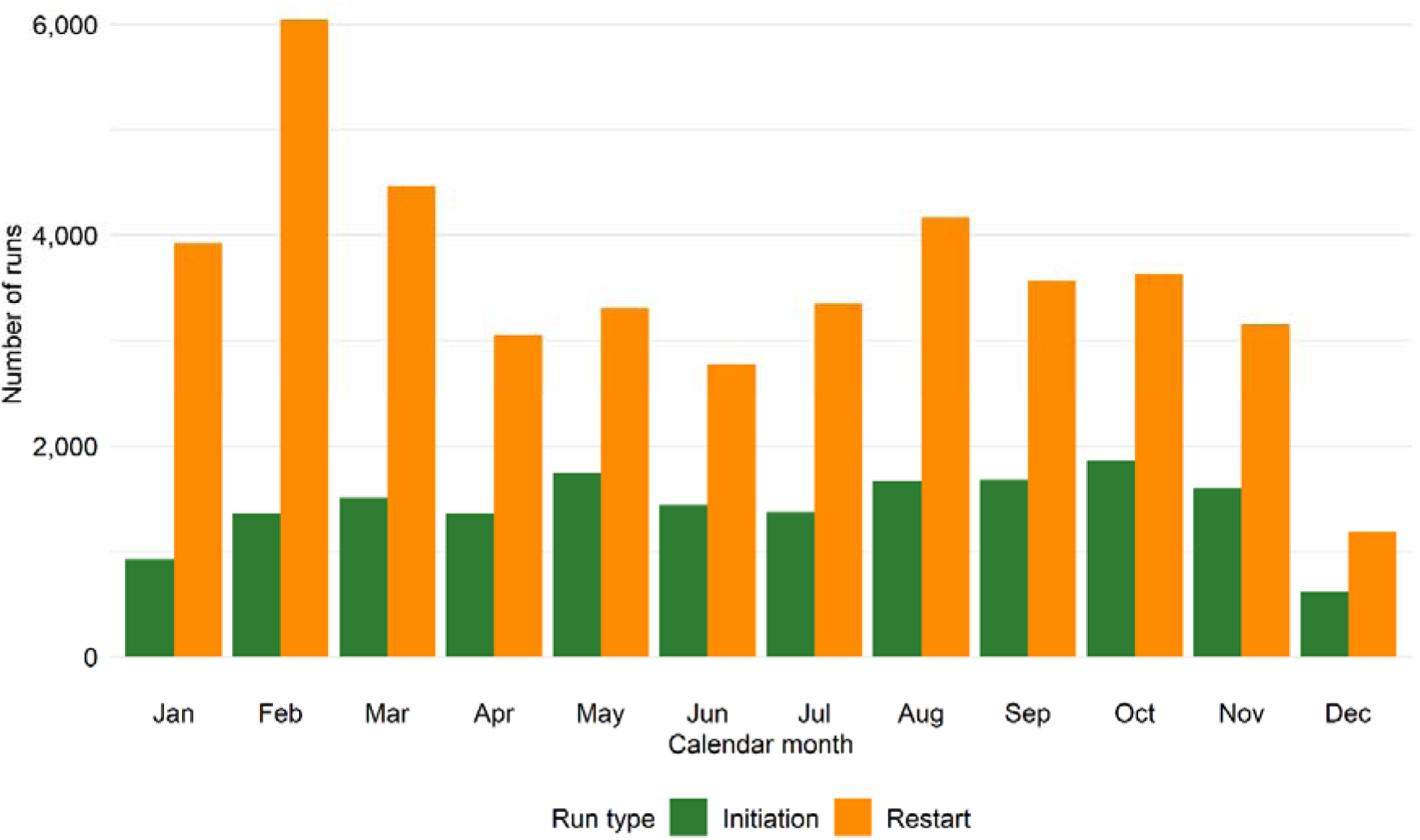
Seasonal patterns in methylphenidate treatment episode initiations by calendar month. Western Cape public health services, 2011-2024.

## Discussion

Our study reinforces the important caveat for all clinicians and caregivers in our large state South African state service-most children and adolescents commencing stimulant medication for ADHD will not remain on it for a meaningful period, despite it being a lifelong condition. Understanding and supporting persistence is the clinical priority. We demonstrate high rates of early patient attrition, frequent re-initiations and strong temporal relationships of re-initiation with the school academic calendar.

Our retention rates for intitiations of 20·5% at 12 months are significantly lower than those described in a recent study by Brikell et al. for children and adolescents in high income countries where 12-month persistence was 65% for children (aged 4-11) and 47% for adolescents (aged 12-17)^44^. A startling 25·4% of first-time methylphenidate prescriptions in our study were never followed by a subsequent dispensing event. This drop-off proportion was higher still for those restarting medication, rising to 29·7%. Our cohort replicated similar findings from persistence literature, namely that gender was not associated with episode duration, but that initiation at an older age was associated with a shorter episode durationsh^8, 44^. Methylphenidate formulation was a complex consideration in our study as access to SR methylphenidate in the state health service was lost in 2022 (prior event in 2008) because of perceived unaffordability. Our study confirms that lack of access to an SR formulation is a significant disadvantage for medication persistence in our setting, in keeping with a well attested literature in this regard^24, 45, 46^.

The most important positive predictors of 12-month persistence with therapy were the frequency with which a patient was seen in the first 3 months of treatment and the incorporation of SR methylphenidate into their treatment. Patients seen at shorter than monthly intervals had a nearly 3-fold increase in odds of maintaining treatment compared to those who saw a clinician at frequencies of longer than a month (OR 2·85 for high visit frequency vs 1·24 for SR methylphenidate in the multivariable model). This is a finding similarly described in two studies in North America, where early contact and dose titration favourably predicted higher script fulfilment rates^20, 47^. The Dundee ADHD Clinical Care Pathway, widely acknowledged as a care exemplar, builds in weekly consultations in the first month of medical therapy intervention^48^. The elements of strong therapeutic alliances are well described in many national ADHD practice guidelines^13–15, 17, 49–51^. Our data do not allow us to interrogate drivers of earlier consultations, which could be both carer and clinician mediated.

Facility transitions occurred in a minority of treatment episodes (7·5%) yet were associated with markedly longer episode durations (8·4 vs 2·6 months). For our state service population, largely reliant on public transport, the ability to reach a service appears to be a more important consideration than clinician continuity. In resource constrained settings where private vehicle ownership is limited, the option of a clinic closer to home would be more desirable than continuity with a specific clinician.

The provision of SR formulation methylphenidate, while not as strongly predictive as early visit frequency in medication persistence, provided consistent positive associations for retention when compared to children who were only prescribed the IR formulation. This is a consistent finding in published literature^46, 52, 53^. In our school systems, the requirement for midday dosing during the school day adds a layer of adherence complexity, and without this second dose, medication responses can appear inconsistent to both teachers and parents.

Nearly 40% of patients had 3 or more treatment episodes, a pattern of repeated disengagement and return. These are families that return for treatment, implying the perception of benefit from medication but raising the question of what drives the intervening gaps and how service design might shorten them. This cycling pattern is consistent with a model of ADHD as a condition with variable severity across time and context, described by Sonuga-Barke et al and others, where context functions as both mediator and moderator of perceived benefit^54–56^. The peaks and troughs associated with the school academic year and vacations may represent crisis responses after periods of “trying without”.

Prescription initiations peak at different points across the school year, and the marked surge in re-initiations following the mid-year and end-of-year vacations points to academic demands as the external factor driving re-engagement rather than any adherence pause related to holidays or personal need. Our 90-day treatment gap criterion exceeds the duration of school holidays, meaning these re-initiations constitute genuine treatment restarts. While local and international literature notes a dip in prescribing during school holidays and an increase before exams, our cohort shows increased restart activity at the start of each half of the academic year, consistent with uptake driven by the resumption of school academic demands^29, 57–59^.

Our study contains a large dataset of over 420,000 prescriptions for over 65, 000 treatment episodes, over 14 years and 115 facilities. It is derived from a limited-capacity health system with a comprehensive electronic prescription record database. To our knowledge we have no large-scale reference data from LMICs, and our 12-month survival figures start to map core challenges of a medication ecosystem for ADHD in resource-limited settings. It offers a valuable window into the relationship between a complex condition and the way in which health workers administer stimulant medication and how people access it over time ^34, 60^. Prescription patterns are well differentiated, and we provide evidence of promising avenues to pursue for supporting long term adherence and persistence in ADHD care in an LMIC: early attentive appointments and the provision of SR methylphenidate. We also highlight the phenomenon of school calendar driven stimulant uptake, a phenomenon that deserves broader engagement with families and colleagues in education.

A prescription database of this form is a record of dispensed medication, opaque to the way consultations occurred and the many unique interactions with a health service that enhance or impair access. Additional limitations in the data include the lack of a confirmed diagnosis, no dosing information, the inability to compare clustered prescription fulfilments and clinician assigned visit dates, the circumstances relating to treatment discontinuation and socioeconomic details. The data did not allow for reproduction of either Proportion of Days Covered (PDC) or Medication Possession Ratio (MPR). As patients could contribute multiple restart episodes, these were treated as independent observations; the potential for within-patient correlation across restart episodes was acknowledged as a limitation.

The findings of this study could be used to inform enhanced treatment strategies such as shortened contact periods during commencement, potentially using mental health nurses via telephonic support, and the inclusion of SR methylphenidate as a permanent item in our provincial formulary. The latter option would require sustained advocacy with global pharmaceutical providers to ensure affordable pricing costing for our service. For many of the patients represented in this data, poverty would impair their ability to access services on a monthly basis and we would add our support to a recent request for a review of the regulatory frameworks that governing stimulant scheduling and dispensing to address this barrier^31^.

The phenomenon of the positive deviants in our study warrants further investigation and it is our intention to better understand the drivers of patient attrition. Qualitative research is needed to understand what the factors were that enabled early contact. We intend to investigate the effect of a pragmatic and locally relevant standard of care using simple strategies such as early patient contact and time-efficient access to medication.

## Data Availability

Data does not have approval for sharing.

## Acknowledgements and Disclosures

Ultimate responsibility for data curation and analysis resides with the first author.

MR conceptualised the study, cleaned the dataset and provided initial data analysis and drafting of the text. KD and HMD contributed to early conceptualisation. KD, VR, EL and HMD reviewed the analysis and provided critical review and revision of the manuscript.

## Data availability

The data that support the findings of this study are held by the Western Cape Department of Health and are not publicly available.

## Declaration of AI use

Claude Sonnet 4·6 (Anthropic, PBC) was used between November 2025 and February 2026 to assist with code generation for data cleaning, curation, analysis, table preparation and graphic generation for the statistical package R (Posit Software, PBC, version 4·5.0, 2025-04-11 ucrt using RStudio 2025·09·1), and to identify and retrieve candidate reference literature via an integrated connection to the PubMed database. All AI-generated code and output were reviewed, tested and verified by the authors, and all retrieved references were checked against their original sources.

